# Improving Rectal Tumor Segmentation with Anomaly Fusion Derived from Anatomical Inpainting: A Multicenter Study

**DOI:** 10.1101/2024.10.15.24315517

**Authors:** Lishan Cai, Mohamed A. Abdelatty, Luyi Han, Doenja M. J. Lambregts, Joost van Griethuysen, Eduardo Pooch, Regina G.H. Beets-Tan, Sean Benson, Joren Brunekreef, Jonas Teuwen

**Affiliations:** Department of Radiology, Netherlands Cancer Institute, The Netherlands; GROW School for Oncology and Developmental Biology, Maastricht University Medical Centre, The Netherlands; Department of Diagnostic and Interventional Radiology, Kasr Al-Ainy Hospital, Egypt; Department of Radiology and Nuclear Medicine, Radboud University Medical Centre, The Netherlands; Department of Cardiology, Amsterdam Cardiovascular Sciences, Amsterdam University Medical, Centre, The Netherlands; Department of Radiation Oncology, Netherlands Cancer Institute, The Netherlands; Radboud University Medical Center, Department of Medical Imaging, The Netherlands; University of Amsterdam, Faculty of Science, The Netherlands

## Abstract

Accurate rectal tumor segmentation using magnetic resonance imaging (MRI) is paramount for effective treatment planning. It allows for volumetric and other quantitative tumor assessments, potentially aiding in prognostication and treatment response evaluation. Manual delineation of rectal tumors and surrounding structures is time-consuming and typically. Over the past few years, deep learning has shown strong results in automated tumor segmentation in MRI. Current studies on automated rectal tumor segmentation, however, focus solely on tumoral regions without considering the rectal anatomical entities and often lack a solid multicenter external validation. In this study, we improved rectal tumor segmentation by incorporating anomaly maps derived from anatomical inpainting. This inpainting was implemented using a U-Net-based model trained to reconstruct a healthy rectum and mesorectum from prostate T2-weighted images (T2WI). The rectal anomaly maps were generated from the difference between the original rectal and reconstructed pseudo-healthy slices during inference. The derived anomaly maps were used in the downstream tumor segmentation tasks by fusing them as an additional input channel (AAnnUNet). Alternative methods for integrating rectal anatomical knowledge were evaluated as baselines, including Multi-Target nnUNet (MTnnUNet), which added rectum and mesorectum segmentation as auxiliary tasks, and Multi-Channel nnUNet (MCnnUNet), which utilized rectum and mesorectum masks as an additional input channel. As part of this study, we benchmarked nine models for rectal tumor segmentation on a large multicenter dataset of preoperative T2WI as the baseline and nnUNet outperformed the other eight models on the external dataset. The MTnnUNet demonstrated improvements in both supervised and semi-supervised settings (AI-generated rectum and mesoretum were used) compared to nnUNet, while the MCnnUNet showed benefits only in the semi-supervised setting. Importantly, anomaly maps were strongly associated with tumoral regions, and their integration within AAnnUNet led to the best tumor segmentation results across both settings. The effectiveness of AAnnUNet demonstrated the value of the anomaly maps, indicating a promising direction for improving rectal tumor segmentation and model robustness for multicenter data.

## 1. Introduction

Magnetic Resonance Imaging (MRI) plays a pivotal role in staging rectal cancer and selecting treatment plans, providing valuable information regarding the extent of tumor infiltration within and beyond the bowel wall, and into critical anatomical structures, including perirectal vessels, the mesorectal fascia (MRF), peritoneum, and neighboring pelvic organs (Beets-Tan et al., 2018; MERCURY Study Group, 2007). T2-weighted imaging (T2WI) forms the mainstay of the MRI protocol because of its superior soft tissue contrast to discern the different layers of the rectal wall, mesorectal fat, adjacent vessels, and MRF to allow for detailed local staging (Horvat et al., 2019; Suzuki et al., 2008).

Precisely segmenting the tumor is an important task in rectal cancer management. Tumor segmentations are utilized for several purposes including radiation treatment planning, volumetric analysis, and extraction of imaging biomarkers, which may serve as a basis for prognostication and treatment response evaluation. Manual rectal tumor delineation by experienced radiologists is considered the current gold standard. Nevertheless, it is time-consuming and subject to substantial intra- and inter-observer variation (Hearn et al., 2020; Irving et al., 2016; Trebeschi et al., 2017). Developing an accurate, generalizable, and robust rectal tumor segmentation model can help reduce this variability and assist in standardizing several steps of diagnostic and therapeutic rectal cancer management.

Deep learning (DL) has seen a rapid uptake in several fields, achieving state-of-the-art results in multiple medical image analysis tasks (Razzak et al., 2018). In particular, Convolutional Neural Network-based (CNN) based DL approaches excel in learning image representations from annotated data by using learnable feature extraction filters and sequential convolution, activation, and pooling operations. Among CNN architectures, the U-Net (Ronneberger et al., 2015) and its variants are the most popular architecture in medical image segmentation. Studies (Jian et al., 2018; Knuth et al., 2022; Wang et al., 2018) have explored the ability of U-Net and other 2D CNNs on rectal tumor segmentation based on T2WI. These CNNs achieved a Dice similarity coefficient score (DSC) ranging from 0.59 to 0.84 for tumor segmentation. Although 2D CNNs are less computationally expensive, 3D models can leverage richer context to improve predictions (Mlynarski et al., 2019).

(Hamabe et al., 2022) implemented a 3D U-Net, achieving an averaged DSC of 0.73 (0.60-0.80) over 10-fold cross-validation for rectal tumor segmentation. However, no external validation was done in their study. Besides CNN-based models, transformer-based architectures are also being applied in medical image analysis due to their ability to access long-range semantic information (Xiao et al., 2023). (Li et al., 2023) proposed RTAU-Net, a novel 3D dual path fusion network containing a transformer encoder for extracting global contour information of the tumor. RTAU-Net achieved an averaged DSC of 0.80 and 0.68 in data from two respective medical centers. However, RTAU-Net requires the manual removal of tumor-free slices, which hinders the fully automated implementation of the model. Additionally, RTAU-Net was not compared with the state-of-the-art medical segmentation networks, such as nnUNet (Isensee et al., 2021), a self-configuring implementation of the U-Net architecture, or nnFormer (Zhou et al., 2023), which introduces 3D transformer blocks on top of nnUNet.

Besides rectal tumor delineation, some studies also demonstrated that CNNs can accurately delineate anatomical structures such as rectum and mesorectum or perirectal fat with DSC above 0.90 (DeSilvio et al., 2023; Hamabe et al., 2022; Kim et al., 2019). Automated rectum and mesorectum delineation could potentially improve radiological evaluation. The prognosis for rectal cancer depends on how far the tumor has infiltrated the layers of the rectal wall and the mesorectum, and the successful attainment of negative circumferential resection margins (CRMs) through surgical intervention (Nagtegaal et al., 2004). Additionally, Lee et al. (Lee et al., 2019) demonstrated that a 2D model’s variance in tumor regions can be reduced by 90% by incorporating rectal segmentation on the model’s objective. Integrating rectal anatomical knowledge can provide a more comprehensive representation of the T2WI, allowing the model to learn richer and more nuanced patterns, leading to better performance on unseen data. However, the impact of adding rectal anatomical structures including mesorectum for rectal tumor segmentation has not been investigated in a multi-institutional setting. Also, incorporating rectal anatomical structure has so far been limited to adding additional segmentation tasks as presented in (Lee et al., 2019).

Unlike for medical challenges with public datasets like brain tumor segmentation (Menze et al., 2015) or clinically significant prostate lesion segmentation (PICAI) (Saha et al., 2023), there is no large multicenter publicly available MRI dataset for rectal cancer studies. This makes it difficult to benchmark different models. An extensive external validation study with multicenter data is highly desirable to compare different deep learning segmentation approaches.

In this article, we had the following contributions:

- We developed and evaluated a rectal tumor segmentation model incorporating anomaly maps from anatomical inpainting, showing improved performance.
- To generate the anomaly maps, we proposed and evaluated a novel end-to-end rectal anatomical inpainting model, trained exclusively on prostate T2WI, to detect and highlight anomalous areas in rectal T2WI. The model trained on prostate T2WI was applied to rectal T2WI, challenging traditional domain-specific practices and demonstrating the potential for transfer learning. The derived rectal anomaly maps can also be integrated into other clinical downstream tasks.
- As part of our study, we benchmarked nine 3D deep learning models for rectal tumor segmentation on a large multicenter dataset of T2WI.
- We developed and evaluated a 3D deep learning model specifically to segment rectal anatomical structures, including rectum and mesorectum.
- We explored different strategies for incorporating rectal anatomical information into rectal tumor segmentation, including the integration of anomaly maps derived from the inpainting model, adding rectal structures as additional tasks, and utilizing them as prior knowledge.
- We released the rectum and mesorectum masks of 100 prostate T2WIs from PICAI dataset, annotated by radiologist.
- We uploaded the model weights for rectum and mesorectum segmentation, as well as the weights for MTnnUNet, MCnnUNet, and AAnnUNet.

## 2 Related Work

### 2.1 Reconstruction-based Medical Anomaly Detection

Supervised learning requires a substantial amount of reliably labeled data, which is often difficult to collect in medical imaging. Therefore, methods requiring partly labeled data (semi-supervised) or no labeling (unsupervised methods) have attracted increased attention. Anomaly detection is a method that can use semi-supervised or unsupervised techniques to address medical imaging tasks such as segmentation. Generative models are frequently used in the field of anomaly detection due to their effectiveness. The mode is often trained to reconstruct images from a specific data distribution (e.g., normal tissue). These models, when confronted with images outside of this distribution (such as those containing tumors), often struggle to accurately reconstruct the anomalous regions, resulting in higher reconstruction errors in those areas Generative adversarial networks (GANs) (Goodfellow et al., 2014), Auto-Encoders (AE), and their variants including Variational AE (VAE) (Kingma and Welling, 2022) and Vector Quantized VAE (VQ-VAE) (van den Oord et al., 2017) were proven to be promising (Baur et al., 2021; Chen et al., 2020; Pinaya et al., 2022) in this field. However, these methods have presented several challenges. They struggle to learn and represent healthy anatomical structures when using full image resolution. Additionally, when reconstructing the entire image, the highlighted regions might not exclusively correspond to diseased areas. As a result, the final anomaly maps may become more sensitive to other artifacts. Finally, if it is in the self-supervised setting where healthy or normal samples are not available for training, these methods can sometimes can successfully reconstruct anomalies due to a high generalization capacity (Gong et al., 2019).

#### 2.2.1 Partial Image Reconstruction via Inpainting for Medical Anomaly Detection

Instead of image-to-image reconstruction methods, inpainting focuses on filling in missing or occluded parts of an image by leveraging the surrounding context. The masks used for inpainting are generally independent of the dataset. (Nguyen et al., 2021) implemented an inpainting-based brain tumor segmentation pipeline for T1-weighted MRI, where anomalous regions were determined by identifying areas of highest reconstruction loss. With prior anatomical knowledge, inpainting can focus solely on high-risk regions, reducing the impact from the background. (Yeganeh et al., 2022) developed an anatomy-aware masking strategy for inpainting to effectively aid the model in learning the shape representation of the organs of interest. (Woo et al., 2024) have proposed a UNet-based model for detecting bone lesions in knee MR images through reconstruction via inpainting and demonstrated that detected anomalies can be further utilized for segmentation. The core idea of anomaly detection through anatomical inpainting involves masking the region of interest (ROI), typically covering the relevant anatomical structures. The inpainting model is trained to fill in the masked area without potential anomalies. The discrepancy between the original and reconstructed images is then utilized to identify anomalies.

In this study, the anatomical structures strongly associated with rectal cancer, including the rectum and mesorectum (fatty tissue surrounding the rectum), were masked, and the inpainting model was trained using prostate T2WI images from the PICAI dataset (Saha et al., 2023). Despite the different fields of view between prostate T2WI and rectal T2WI, they overlap in anatomical structures. Most prostate T2WI contains a healthy rectum and mesorectum, ensuring the inpainting model was trained to learn the distribution of the healthy tissues. The inferred reconstructed rectal T2WI slices can then be used to generate anomaly maps, highlighting potentially tumoral areas.

## 3. Methods

### 3.1 Rectal Anomaly Detection with Masked T2WI using Anatomical Inpainting

The overall pipeline, inspired by (Han et al., 2024, 2023), for detecting rectal anomalies is shown in Figure 2. Prostate T2WI with masked rectum and mesorectum were used to train the model for their reconstruction. The inpainting model was adapted from (Han et al., 2024), an end-to-end MRI sequence generation framework. The framework consists of two stages. In the first stage, only the reconstruction loss is optimized for the encoder and generator. In the second stage, both adversarial loss and cycle-consistent loss are incorporated in addition to the reconstruction loss, with the optimization applied to the encoder, generator, and discriminator. The training of the anatomical inpainting was based on 2D slices. The inpainting model contains an encoder ***E*** and a decoder ***G***. A masked (rectum and mesorectum) 2D T2WI slice *X*, can be compressed by ***E*** into a latent space *z* = ***E***(*X*) and ***G*** can reconstruct the original slice from the latent representation *z*. The skip connections were added to recover fine-grained details. To enforce similarity between the generated slices and the actual slices, a supervised reconstruction loss is used:

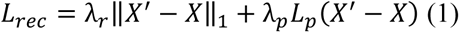

where *X* is the original slice, and *X*^′^ = ***G***(***E***(*X*)) is the restored image. ‖⋅‖_1_ is the *L*_1_ loss and *L*_*p*_ is the perceptual loss from pre-trained VGG19, which involves comparing high-level features (not just pixel values) from both the generated and reference images (Johnson et al., 2016). Instead of measuring raw pixel differences, it evaluates how similar the images are in terms of their content and style, based on features extracted from different layers of the VGG19 network. λ_*r*_ and λ_*p*_ are weight factors, for which the respective values 10 and 0.01 were chosen empirically.

**Figure 1:**
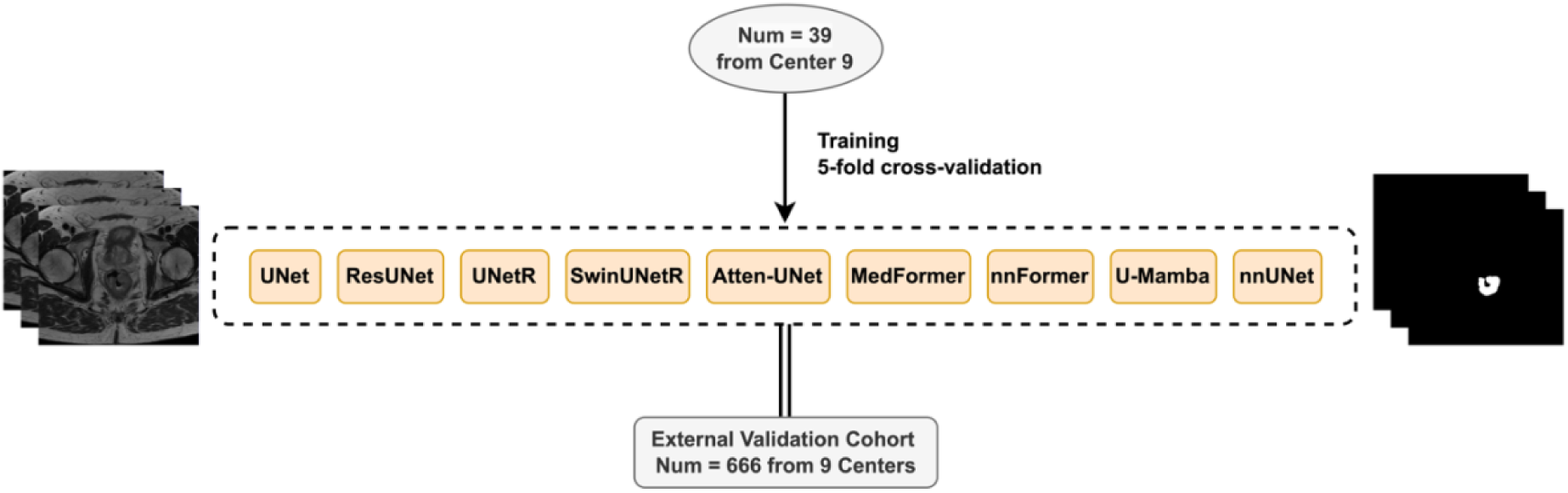
The rectal tumor segmentation performances of various 3D DL models were compared. All the models were trained with training cohort 1 including 39 samples from Center 9 only and externally tested with 666 samples from 9 centers.

**Figure 2:**
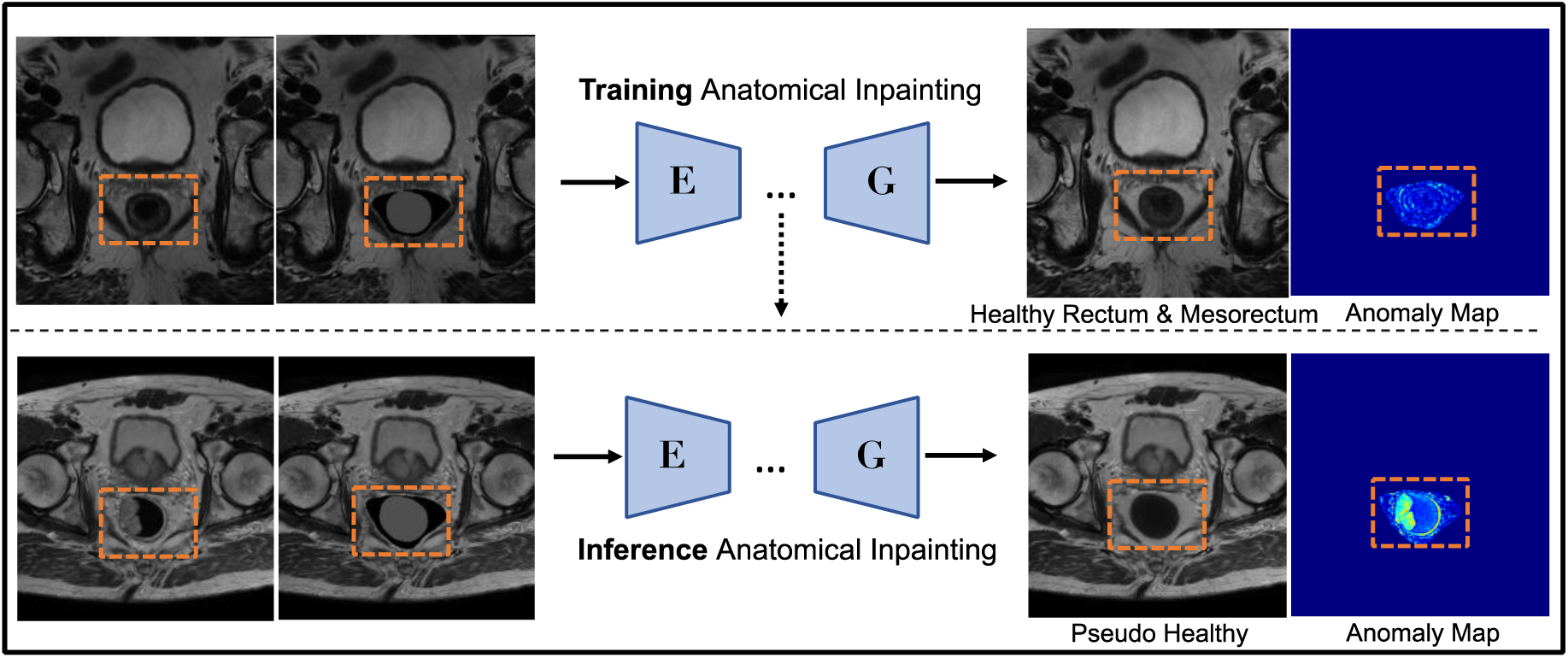
The anatomical inpainting workflow. The inpainting model containing an encoder **E** and a decoder **G** was trained using prostate T2WI with a healthy rectum and mesorectum. The trained model was inferred across the rectal dataset to generate a reconstructed pseudo healthy rectum and mesorectum. The difference between the reconstructed and the original image is the anomaly map where tumoral regions were highlighted.

For the second stage of the training, the adversarial loss and cycle-consistent loss (Zhu et al., 2017) were added on top of the reconstruction loss to ensure that the inpainted images were both realistic and consistent with the original images. The adversarial loss helps to ensure that the completed regions look realistic and blend seamlessly with the surrounding areas and the cycle-consistent loss focuses on preserving the original structure by ensuring the inpainted image can be accurately reconstructed back to the original.

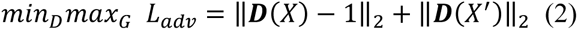

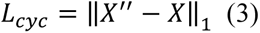

where *X*^′′^ = ***G***(***E***(*X*^′^)) and ‖⋅‖_2_ is the *L*_2_ loss and ***D*** is the discriminator. The anomaly maps are then defined as the absolute differences between the reconstructed slice and the original slice,

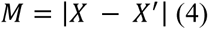

Let *I* be an image with intensity values. The normalization scheme involved the following steps:

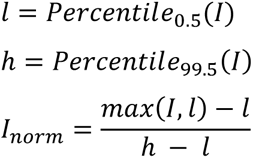

First Compute the 0.5th percentile (*l*) and the 99.5th percentile (ℎ) of the intensity values in the image and normalize the intensity values in the image using the computed percentiles. The rectum was masked with a value of 0, while the mesorectum was masked with a value of 0.5. The model input patch size was (384, 384), with a batch size of 1. We used the AdamW optimizer to train the network (both stages) with β 0.9 and 0.95, an initial learning rate of 0.0001, a weight decay factor of 0.05, and following a polynomial decay. The model was implemented in PyTorch (Torch version 2.1.2) and the training was conducted on an NVIDIA RTX A6000 GPU.

### 3.2 Study Design

To train the inpainting model, 100 prostate T2WI with manually segmented rectum and mesorectum masks were split into 80 for training and 20 for internal validation. The model was additionally tested on 200 prostate T2WIs and the entire rectal dataset, comprising 705 T2WI. However, because annotating rectal structures is labor-intensive, only 180 rectal randomly selected samples have radiologist-annotated masks for the rectum and mesorectum. To address this, a nnUNet, defined as anatomy nnUNet, was trained specifically to segment the rectum and mesorectum using a dataset of 39 samples (training cohort 1) from a single center, which had the highest number of manually annotated rectum and mesorectum masks among nine centers. Some studies have demonstrated that CNNs can accurately delineate anatomical structures such as rectum and mesorectum or perirectal fat with DSC above 0.90 (DeSilvio et al., 2023; Hamabe et al., 2022; Kim et al., 2019). The model was then evaluated on 141 external samples, see Figure 3. This nnUNet was then used to infer all the rectum and mesorectum masks across the whole rectal dataset. The predicted rectum and mesorectum masks were defined as AI-generated pseudo rectum and mesorectum masks.

**Figure 3:**
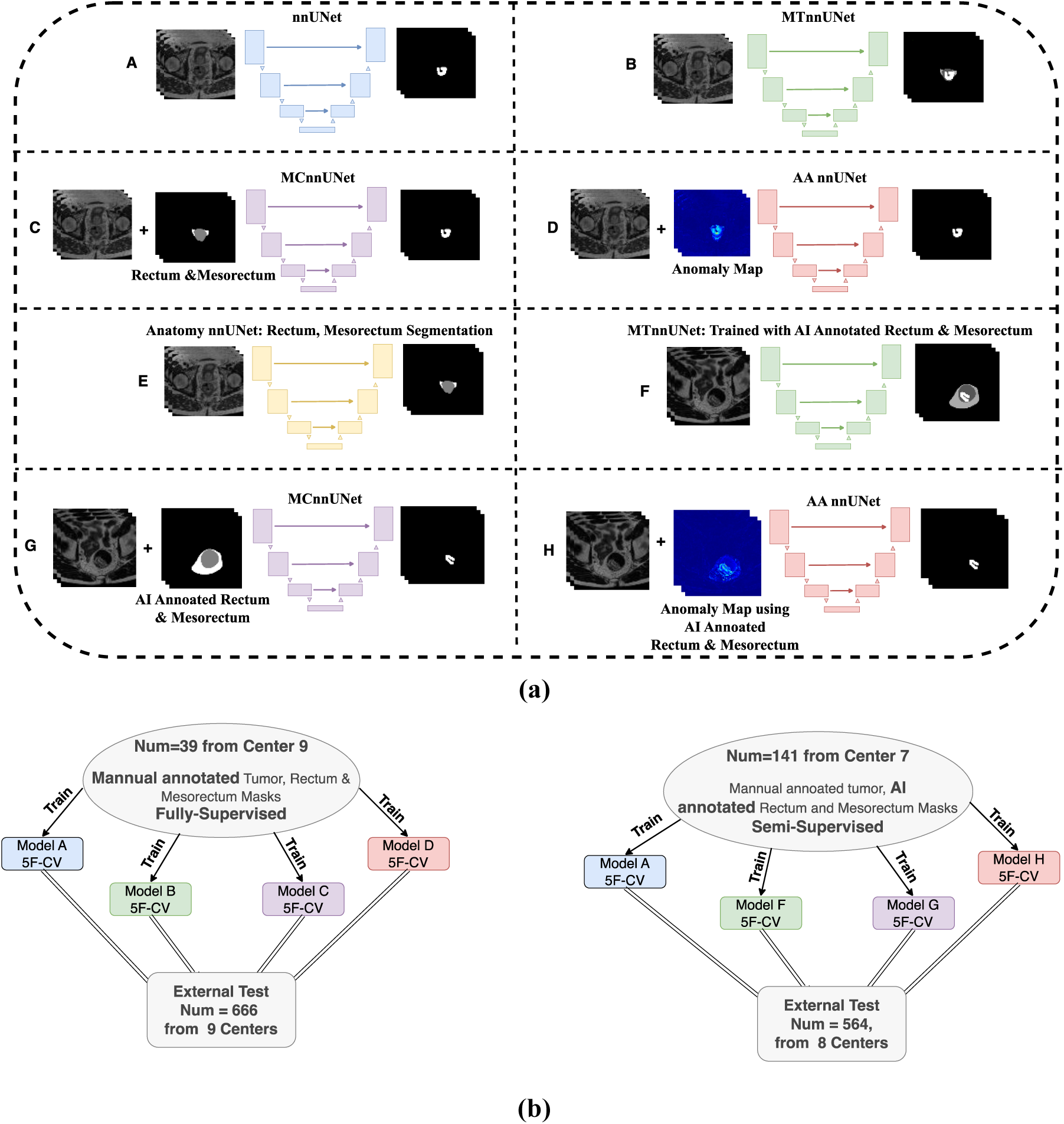
(**a)** Model definition. A: a nnUNet for tumor segmentation only; B: MTnnUNet: multi-target nnUNet, segmenting tumor, rectum, and mesorectum simultaneously, C: MCnnUNet: multi-channel nnUNet with rectum and mesorectum masks together with T2WI as input, D: AAnnUNet: Anomaly-Aware nnUNet, with anomaly maps derived from anatomical inpainting as additional input E: Anatomy nnUNet, to segment rectum and mesorectum. F: MTnnUNet but trained with AI-generated rectum and mesorectum, G: MCnnUNet but using AI-generated rectum and mesorectum, H: AAnnUNet but using AI-generated rectum and mesorectum **(b)** The training scheme for different models. 5F-CV: 5-fold cross-validation. Models A (A-H) are defined in (**a**).

We incorporated anomaly maps generated by the inpainting model into downstream rectal tumor segmentation tasks by adding them as an additional input for nnUNet, called Anomaly-Aware nnUNet (AAnnUNet). We compared this approach with other strategies for integrating anatomical knowledge into tumor segmentation, including Multi-Target nnUNet (MTnnUNet) and Multi-Channel nnUNet (MCnnUNet). MTnnUNet added rectum and mesorectum segmentation as auxiliary tasks, and Multi-Channel nnUNet (MCnnUNet) utilized rectum and mesorectum masks as additional input channels. First, we established a baseline for rectal tumor segmentation by comparing the performance of nine 3D deep learning models, see Figure 1, including UNet (Çiçek et al., 2016), ResUNet (Diakogiannis et al., 2020), UNetR (Hatamizadeh et al., 2022b), SwinUNetR (Hatamizadeh et al., 2022a), AttentionUNet (Atten-UNet) (Oktay et al., 2018), MedFormer (Gao et al., 2023), nnFormer (Zhou et al., 2023), U-Mamba (bot) (Ma et al., 2024) and nnUNet (Isensee et al., 2021). All the models underwent training using training cohort 1 with a 5-fold cross-validation. Subsequently, models were externally tested on the remaining 666 samples from nine centers. MTnnUNet, MCnnUNet, and AAnnUNet were trained on training cohort 1 using 5-fold cross-validation and tested on 666 samples from 9 centers. For the inference of MCnnUNet and AAnnUNet, AI-generated pseudo rectum and mesorectum were used.

With AI-generated pseudo anatomical structures, MTnnUNet, MCnnUNet, and AAnnUNet were also trained on a larger cohort comprising 141 samples (training cohort 2) using 5-fold cross-validation and tested on 564 samples from eight centers. Instead of relying on ground truth rectum and mesorectum masks, AI-generated annotations were employed in training. This method merged AI-labeled anatomical structures with manually labeled tumors, indicating a semi-supervised learning approach.

### 3.3 Segmentation Models

1. **UNet** (Çiçek et al., 2016), extends the previous u-net architecture from Ronneberger et al., 2015 (Ronneberger et al., 2015) by replacing all 2D operations with their 3D counterparts.
2. **ResUNet** (Diakogiannis et al., 2020), is a modified version of UNet. It replaces double convolution layers of UNet with residual blocks from ResNet (He et al., 2016), incorporating shortcut connections for faster convergence. This adaptation works in both 2D and 3D settings, enhancing performance in capturing complex patterns.
3. **UNETR** (Hatamizadeh et al., 2022b), adopts a ViT-inspired encoder and employs a CNN decoder for 3D image segmentation. The images are initially divided into patches, which are linearly transformed into token embeddings. These tokens undergo processing through a self-attention block, akin to ViT. To manage the quadratic complexity of self-attention, the patch size is set to be relatively large (16) to prevent overly long sequence lengths.
4. **SwinUNETR** (Hatamizadeh et al., 2022a), reformulates the segmentation task as a sequence-to-sequence prediction using a Swin Transformer as the encoder. The encoder is then connected to a Fully Convolutional Neural Network (FCNN)-based decoder through skip connections.
5. **Attention UNet** (Oktay et al., 2018), introduces an attention-gating module to UNet to enhance its ability to suppress irrelevant regions and highlight salient features crucial for a given task.
6. **nnFormer** (Zhou et al., 2023), is a 3D transformer for volumetric medical image segmentation nnFormer combines interleaved convolution and self-attention operations. It introduces a special self-attention mechanism to understand both local and global aspects of the image volume. To improve efficiency, it also uses skip attention instead of the usual concatenation or summation operations.
7. **MedFormer** (Gao et al., 2023), is a transformer-based designed to handle scalable 3D medical image segmentation, including three crucial components: a beneficial inductive bias, hierarchical modeling using linear-complexity attention, and multi-scale feature fusion that combines spatial and semantic information globally. MedFormer can learn from both small and large scale data without pre-training.
8. **U-Mamba** (Ma et al., 2024), is inspired by the State Space Sequence Models (SSMs) (Gu et al., 2021), which are known for their ability to handle long sequences. The model is designed specifically for biomedical image segmentation with the hybrid CNN-SSM block that integrates the local feature extraction power of convolutional layers with the abilities of SSMs to capture the long-range dependency.
9. **nnUNet** (Isensee et al., 2021), is a self-configuring framework for medical image segmentation. It utilizes UNet as its architecture but offers a specialized preprocessing, training technique, and hyper-parameter configuration. nnUNet achieves state-of-the-art performance on several medical image segmentation challenges with a relatively simple architectural design.
10. **Anatomy nnUNet**, is the nnUNet trained to segment rectal-related anatomical structures including the rectum and mesorectum.
11. **MTnnUNet**, is the nnUNet trained to segment rectum, mesorectum, and rectal tumors.
12. **MCnnUNet**, is the nnUNet trained to segment rectal tumors with rectum and mesorectum masks as additional input channels.
13. **AAnnUNet**, is the nnUNet trained to segment rectal tumors with anomaly maps *M* derived from anatomical inpainting.

For rectal tumor segmentation, the imaging preprocessing approach was adopted from nnUNet, which included ZScoreNormalization for standardizing intensities, uniform resampling of all images, and a cropping process. All the segmentation models were implemented in PyTorch (Torch version 2.1.2) and trained on an NVIDIA A6000 GPU with randomly initialized weights without transfer learning. The batch size was set to 2 and the models were trained for 1000 epochs with the SGD optimizer. The loss function is the sum of the cross-entropy and Dice loss. During inference, predictions were obtained by averaging the outputs of each model resulting from the 5-fold cross-validation procedure.

### 3.4 Evaluation Metrics and Statistical Analysis

Statistical analysis was conducted in Python (version 3.9) with the SciPy package (version 1.13.1). To measure the performance of image reconstruction, Structural Similarity Index Measurement (SSIM), Peak Signal-to-NoiseRatio (PSNR) were used. To measure the segmentation performance, the Dice Similarity Coefficient score (DSC) and 95% Hausdorff Distance (HD) were utilized on both cross-validation and external tests. The characteristic differences of cohorts were compared by the Kruskal-Wallis test. The Mann–Whitney U-test was used to compare the difference of indicators among different methods. The model performance differences were calculated using the paired sample t-test. All statistical analyses were two-sided and p-values below 0.05 were regarded as statistically significant. 95% confidence intervals were generated using the bootstrap method with 10,000 replications.

## 4. Results

### 4.1 Dataset and Patient Characteristics

As a part of a previous institutional review board approved multicenter study project (Bogveradze et al., 2022; Cai et al., 2024; El Khababi et al., 2023; “ESGAR 2020 Book of Abstracts,” 2020; Schurink et al., 2023, 2022) the clinical and imaging data from 1426 patients with biopsy-proven rectal cancer were retrospectively collected from ten medical centers between 2011 and 2018. The original study was conducted in accordance with the Declaration of Helsinki and informed consent was waived due to the retrospective nature of the study. For the current study, the baseline staging T2WI of 705 patients (from 9 centers) was selected from this previous dataset. Patients were excluded if they had one of the following properties: non-diagnostic image quality issues, multiple tumors in the field of view, abscesses surrounding the tumor, unavailability of pre-treatment T2WI, or the unavailability of axial images. To train the anatomical inpainting model for reconstructing the healthy rectum and mesorectum, 300 samples with healthy rectum and mesorectum were randomly selected from the PICAI dataset.

Table 1 shows the patient characteristics of all the selective patients including age, gender, clinical T and N staging, tumor location, and EMVI status.

**Table 1.** Summary of patient demographic and clinical characteristics of the multicenter dataset.

|  |  | All | Training Cohort 1 | Training Cohort 2 | p-value |
| --- | --- | --- | --- | --- | --- |
| Age (median, range) |  | 65 (26-88) | 65 (39-82) | 66 (44-87) | 0.75*, 0.17* |
| Sex | Female | 247 (35%) | 12 (31%) | 40 (28%) | 0.69**, 0.08** |
|  | Male | 458 (65%) | 27 (69%) | 101 (72%) |  |
| cT | 1-2 | 87 (12%) | 3 (8%) | 19 (13%) | 0.58*, 0.24* |
|  | 3 | 519 (74%) | 34 (87%) | 107 (76%) |  |
|  | 4 | 99 (13%) | 2 (5%) | 15 (11%) |  |
| cN | 0 | 268 (38%) | 14 (36%) | 67 (48%) | 0.52*, 0.001* |
|  | 1 | 264 (37 %) | 13 (33%) | 54 (38%) |  |
|  | 2 | 173 (25%) | 12 (31%) | 20 (14%) |  |
| Location | Low | 432 (61%) | 25 (64%) | 93 (66%) | 0.45*, 0.65* |
|  | Middle | 236(34%) | 14 (36%) | 43 (30%) |  |
|  | High | 37 (5%) | 0 (0%) | 5 (4%) |  |
| EMVI | EMVI + | 274 (39%) | 18 (46%) | 38 (27%) | 0.43**, 0.002** |
|  | EMVI - | 431 (61%) | 21 (54%) | 103 (73%) |  |
| Total |  | 705 | 39 | 141 |  |

Values in age parentheses are the minimum and maximum. Values in parentheses of other items are the percentages. cT, baseline clinical T staging. cN, baseline clinical N staging. Location, tumor location. EMVI+: EMVI positive. EMVI-, EMVI negative.
\*, p values were calculated using the Kruskal-Wallis test between the training cohort, training cohort2 and the external dataset.
\*\*, p values were calculated using the Chi-square test between the training cohort1, training cohort2 and the external dataset.

### 4.2 Ground Truth Segmentation

The ground truth segmentation labels were annotated by a GIT radiologist (M.A.A) with 6-7 years of radiological experience in interpreting rectal MRI. Segmentation masks for the rectum and mesorectum were annotated using 180 randomly selected rectal cases and 100 prostate T2W images. Specifically, the entire rectum was annotated, including its lumen from the anorectal junction to the recto-sigmoid junction, following the definition of the sigmoid take-off as the upper anatomical landmark of the rectum. The mesorectal fat, enveloped by the mesorectal fascia, was identified as the high T2 signal area surrounding the rectum on all sides, thinner anteriorly than postero-laterally (FACG, 2006; Lambregts et al., 2010). Moving caudally towards the lower rectum, the thickness of the mesorectal fat enveloping the rectal wall decreases due to the gradual tapering of the mesorectum (Lambregts et al., 2010). Tumor segmentation was annotated for all the cases (n = 705), where tumors were labeled as abnormal mural growth within the rectal lumen and extending outside into the mesorectum (Bogveradze et al., 2023; Lambregts et al., 2010). See Table S2 for annotation details.

### 4.3 Rectal Anomaly Detection

According to Table 2, the inpainting model demonstrated superior performance and less variance in reconstructing the rectum and mesorectum in prostate T2WI, with an averaged SSIM (aSSIM) of 86.72 and an averaged PSNR (aPSNR) of 25.87, compared to rectal T2WI, which had an aSSIM of 83.38 and an aPSNR of 23.87. These results aligned with expectations. Prostate T2WI was in-distribution and represented healthy samples of the rectum and mesorectum. Conversely, rectal T2WI with tumor regions were out-of-distribution data and exhibited greater discrepancies from their corresponding pseudo-healthy counterparts. Importantly, we observed that aSSIM, 39.32, and aPSNR, 17.50 of tumoral regions are noticeably lower than the tumor-free regions, indicating effective anomaly detection. The anomaly map can be generated from the differences between original and pseudo-healthy images, see Figure 4. Tumoral regions were highlighted in rectal T2WI slices.

**Figure 4:**
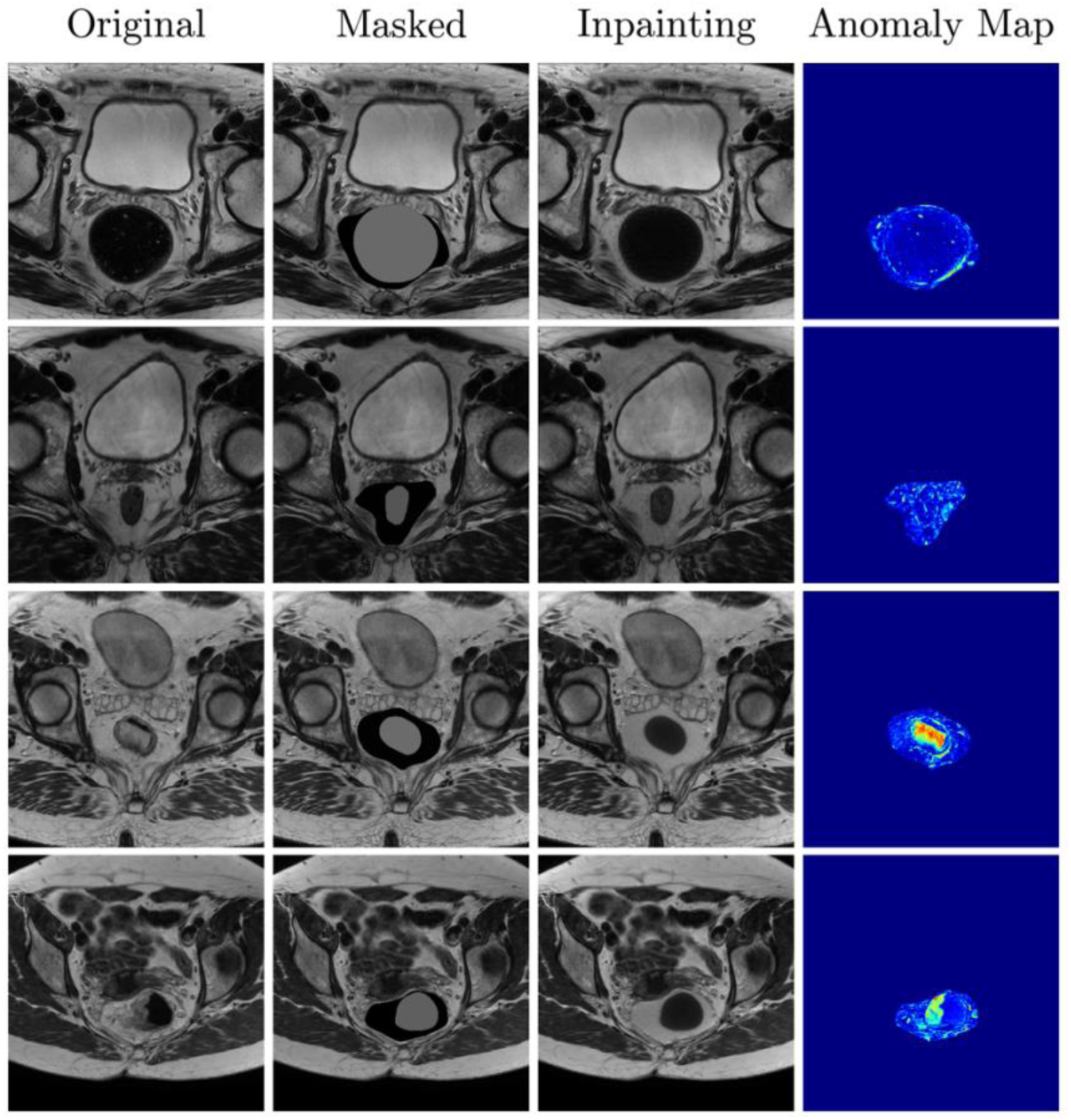
The visualization of anatomical (including rectum and mesorectum) inpainting. The columns from left to right are original T2WI slices, masked slices, inpainted slices, and anomaly map, which is the difference between original and reconstructed slices. The first two rows are prostate T2WI and the last two rows are from rectal T2WI.

**Table 2.**
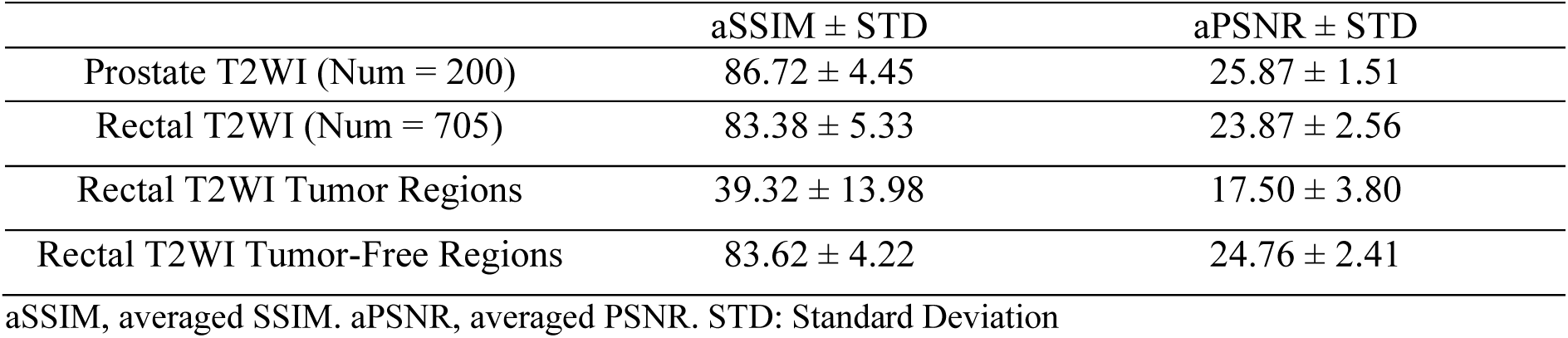
Rectum and Mesorectum inpainting performance in the test data.

### 4.4 The comparison of Nine DL models for Rectal Tumor Segmentation on the external test dataset

In the internal dataset, MedFormer delivered the best overall performance, with an average DSC (aDSC) of 66.3 and a median 95% HD (mHD) of 6.39 mm. U-mamba achieved the highest median DSC (mDSC), while ResUNet excelled in the average 95% HD, see Table S3. However, in the external testing data, as is displayed in Table 3, S4, and Figure 5a, nnUNet achieved the best results, with aDSC 62.8 and aHD 17.28 mm, significantly better than other models. We observed that nnUNet consistently outperformed other models in the external set when the training dataset was increased to 132 from a single center, see Table S10, S11. Additionally, recently proposed transformer-based networks including UNetR, SwinUNet, and nnFormer or the SSMs-inspired architecture like U-Mamba underperformed with respect to CNN-based architectures in external data. We also compared the size of trainable parameters and Floating Point Operations (FLOPs) for each model, see Figure 5b. In summary, nnUNet demonstrated the highest DSC despite having relatively few model parameters and a low number of FLOPs. From Figure 6, nnUNet showed better prediction performance in identifying rectal tumor regions across various centers when using multi-center T2WI. nnUNet exhibited a significantly reduced false positive voxel classification in these displaying cases, especially in the last row of Figure 6, which was a tumor-free slice.

**Figure 5:**
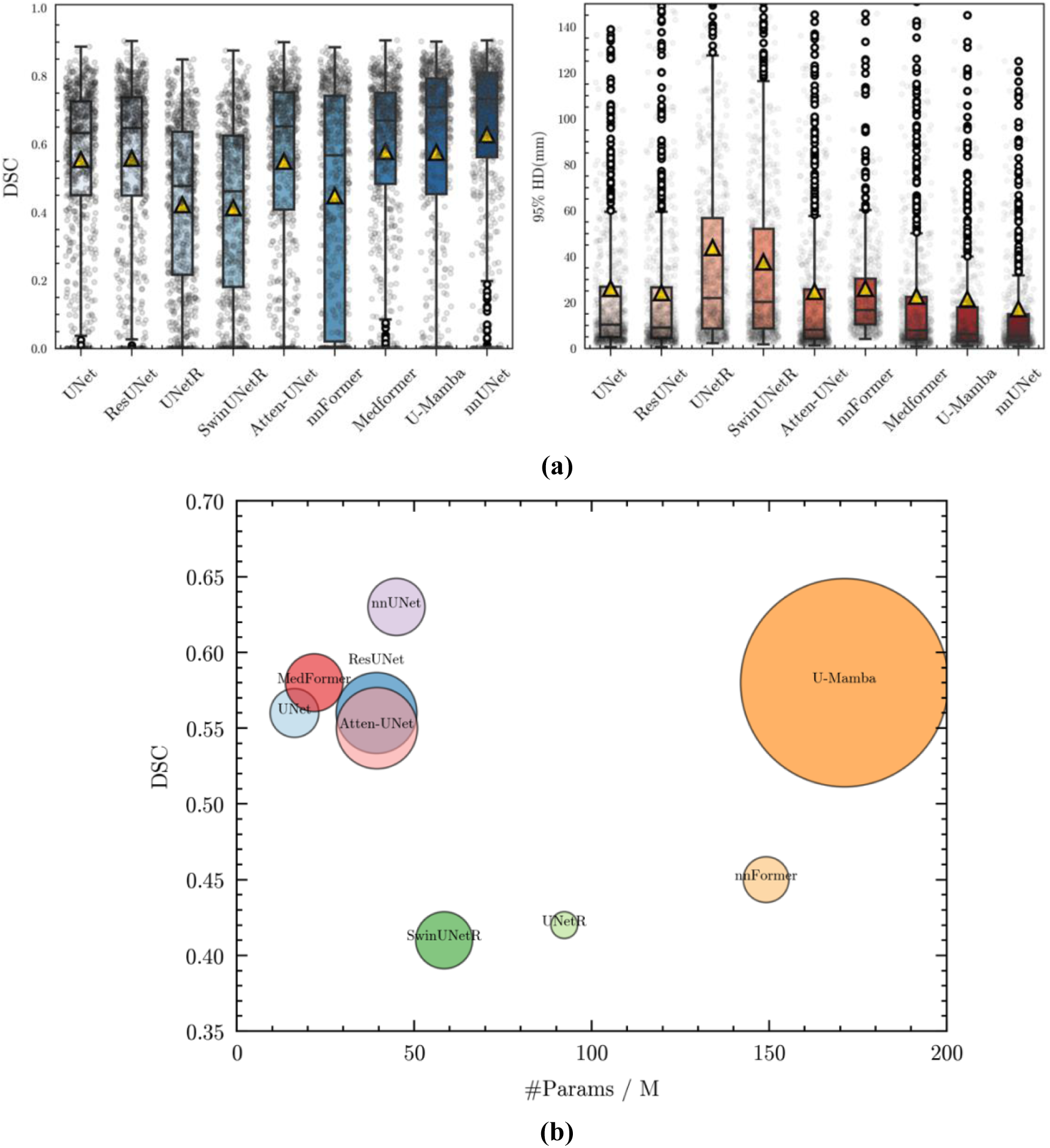
**(a)** The DSC and 95% HD boxplots of various DL models were compared. Yellow triangle denotes the mean DSC. **(b)** DSC v.s. number of trainable parameters v.s. Flops. y-axis: DSC, x-axis: number of parameters/M, bubble size/number under model: the Flops/G. Flops: Floating Point Operations Per Second.

**Figure 6:**
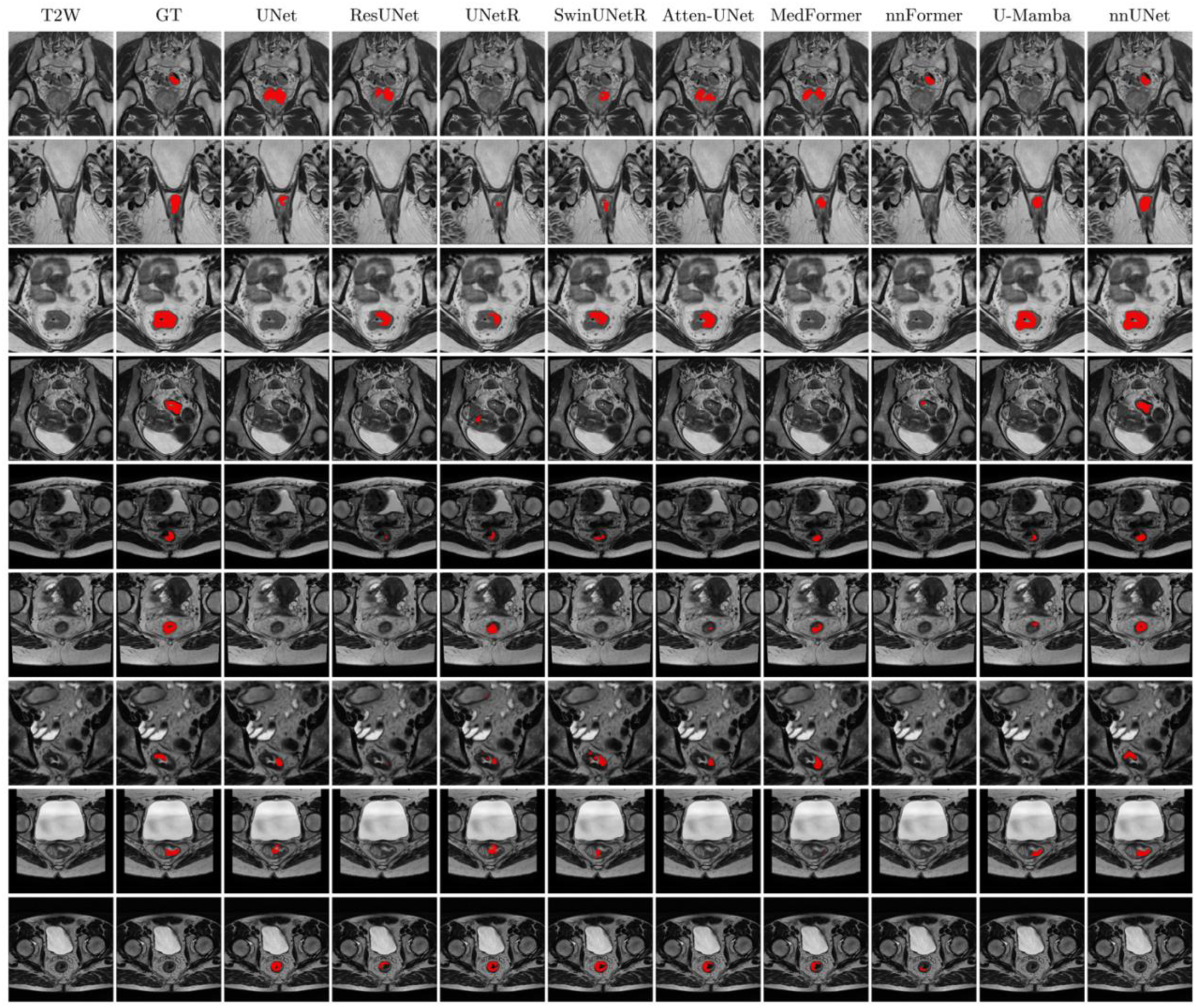
The visualization of the segmentation performance of all nine DL models using T2WI. Each row is a different sample from the external set. The columns from left to right are original T2WI, GT: ground truth, tumor prediction masks from UNet, ResUNet, UNetR, SwinUNetR, Atten-UNet, MedFormer, nnFormer, U-Mamba, nnUNet.

**Table 3.** Comparison of various models on rectal tumor segmentation in the external test (Num = 666, 9 centers)

| Network | UNet | ResUNet | UNetR | SwinUNetR | Atten-UNet | MedFormer | nnFormer | U-Mamba | nnUNet |
| --- | --- | --- | --- | --- | --- | --- | --- | --- | --- |
| aDSC (%) | 55.5<br>(53.7, 57.2) | 55.8<br>(54.0, 57.7) | 42.2<br>(40.4, 44.1) | 41.3<br>(39.4, 43.3) | 55.0<br>(53.0, 57.0) | 57.9<br>(56.0, 59.8) | 44.8<br>(42.4, 47.3) | 57.6<br>(55.4, 60.0) | **62.8**<br>**(60.7, 64.8)** |
| mDSC (%) | 63.2<br>(61.8, 64.7) | 64.8<br>(63.3, 66.3) | 47.8<br>(44.8, 50.7) | 46.2<br>(42.3, 50.1) | 65.2<br>(63.1, 67.2) | 67.0<br>(65.3, 68.7) | 56.7<br>(51.6, 61.8) | 70.9<br>(69.2, 72.6) | **73.2**<br>**(71.8, 74.7)** |
| aHD (mm) | 26.04<br>(22.85, 29.23) | 24.31<br>(21.46, 27.16) | 43.8<br>(39.5, 48.2) | 37.65<br>(34.23, 41.07) | 24.73<br>(21.51, 27.95) | 22.74<br>(19.77, 25.70) | 26.90<br>(23.39, 30.42) | 21.52<br>(17.60, 25.45) | **17.28**<br>**(14.63, 19.94)** |
| mHD (mm) | 10.40<br>(9.32, 11.49) | 9.39<br>(8.39, 10.40) | 22.0<br>(18.4, 25.7) | 20.66<br>(18.04, 23.27) | 8.69<br>(7.48, 9.91) | 8.15<br>(6.93, 9.32) | 16.13<br>(13.41, 18.84) | 7.45<br>(6.54, 8.36) | **6.36**<br>**(5.49, 7.22)** |

aDSC: average Dice Coefficient Similarity. mDSC: median Dice Coefficient Similarity. aHD: average of 95% Hausdorff Distance. mHD: median of 95% Hausdorff Distance. The 95% confidential intervals are presented.

### 4.5 Rectum and Mesorectum Segmentation

nnUNet demonstrated superior performance for rectum segmentation with an aDSC of 0.87 and mHD 10.15 mm, better than the mesorectum segmentation with an aDSC of 0.81, aHD 10.57 mm in the external cohort containing 141 samples see Table S7, Figure S1. This disparity in performance between the rectum and mesorectum is attributed to the rectum’s more regular shape than that of the mesorectum. This is consistent with the findings by (Hamabe et al., 2022). Overall, the segmentation tasks for rectal anatomical structures were significantly easier than for tumors.

### 4.6 MTnnUNet, MCnnUNet and AAnnUNet

From Table 4 and Figure 7a, MTnnUNet and AAnnUNet significantly outperformed both nnUNet and MCnnUNet. Even though MCnnUNet showed the best results in the internal validation set (see Table S8), it exhibited the lowest aDSC and aHD in the external test. The reason could be that MCnnUNet depends more on prior anatomical knowledge and there was substantial inconsistency of anatomical structures in training and inference. MCnnUNet was initially trained with ground truth anatomical structure inputs but utilized AI-annotated structures during inference, causing a performance drop in the external test set. Unlike MCnnUNet, MTnnUNet required anatomical knowledge only during training and did not rely on it during inference. Although AAnnUNet, fusing anomaly maps with highlighted tumoral regions, also relied on the quality of rectum and mesorectum masks, it yielded slightly better results than MTnnUNet in terms of aDSC. Different from MCnnUNet, which incorporated anatomical masks in a straightforward manner, AAnnUNet was provided with the highlighted anomalous areas derived from healthy distributions. We also noted many false-negative cases, where the predicted tumor mask was empty. To address this issue, we combined MTnnUNet and AAnnUNet using a union ensemble approach. This ensemble method improved performance by 3% in aDSC but did not enhance HD, as the union approach did not reduce false positives. Figure 8 illustrates that AAnnUNet performed well in tumor segmentation for both small and large tumors, highlighting the effectiveness of anomaly fusion. However, when anomaly maps were inadequate, AAnnUNet struggled to detect tumors, as shown in the last row of Figure 8. Additionally, there were cases where all models failed to segment the tumor. However, anomaly maps can signal potential issues with high intensity values, as depicted in Figure 9.

**Figure 7:**
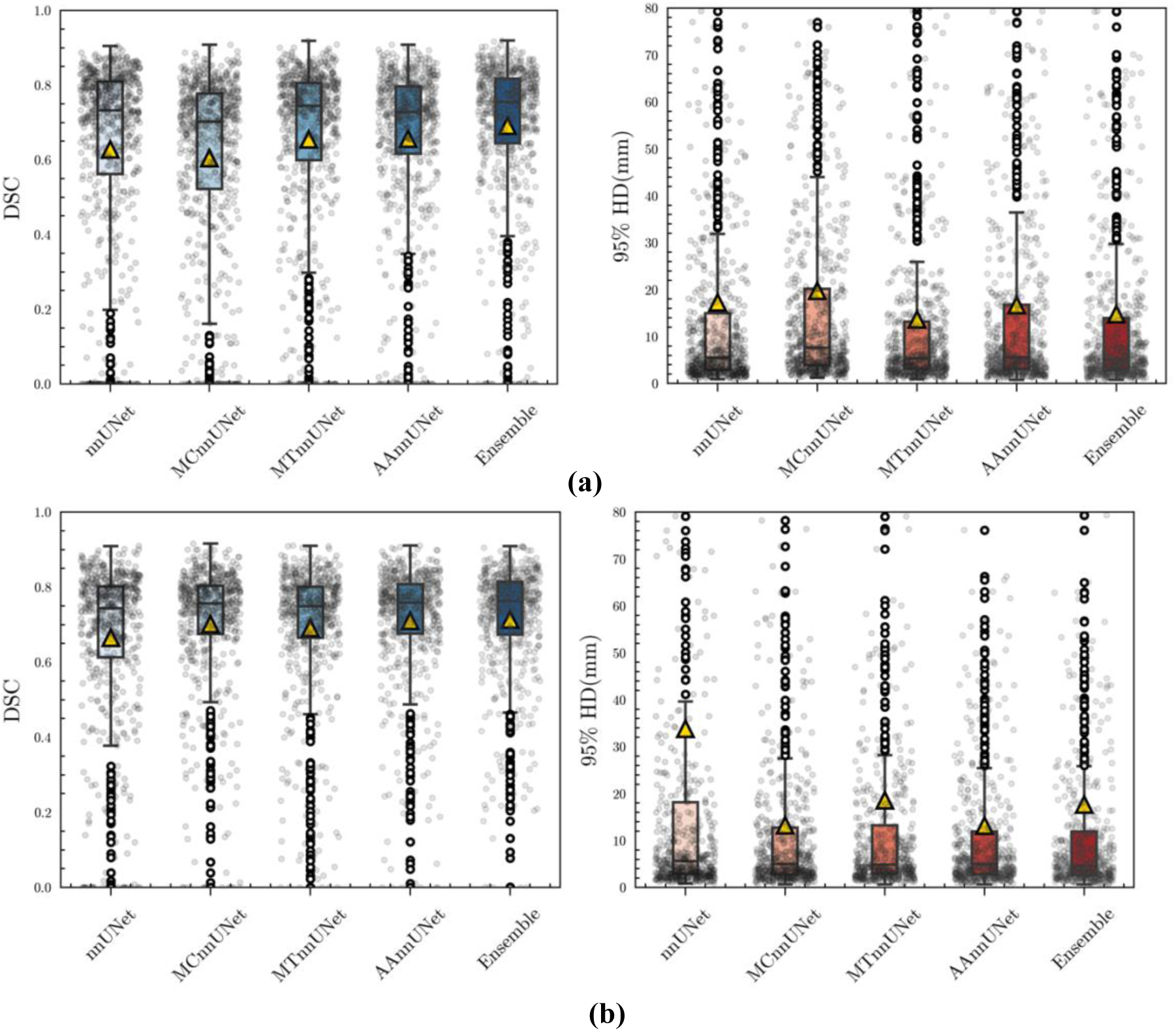
**(a)** The DSC and 95% HD boxplots of nnUNet, MTnnUNet, and MCnnUNet in a fully supervised manner. **(b)** The DSC 95% HD boxplots of nnUNet, MTnnUNet, and MCnnUNet in a semi-supervised manner.

**Figure 8:**
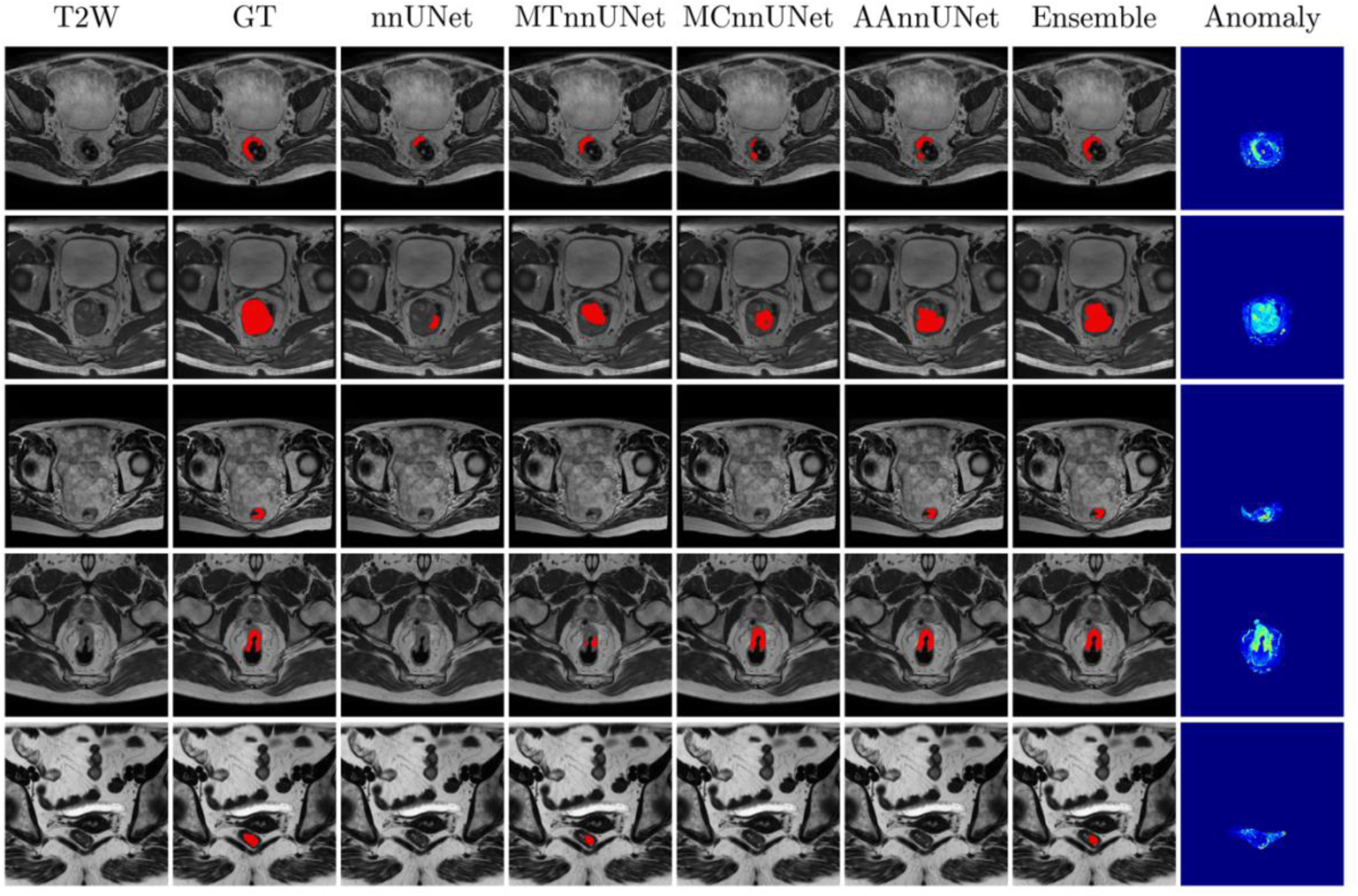
The visualization of the segmentation performance of nnUNet, MTnnUNet, MCnnUNet, AAnnUNet, and Ensemble using T2WI, supervised setting. Each row is a different sample from the external test set. The columns from left to right are original T2WI, ground truth, tumor prediction masks from nnUNet, MTnnUNet, MCnnUNet, AAnnUNet, and Ensemble.

**Figure 9:**
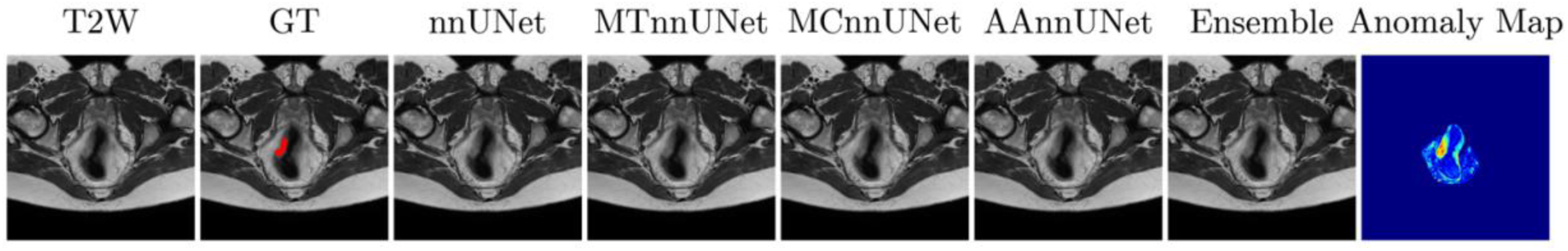
The example where all algorithms failed to locate and segment the rectal tumor, but the anomaly map highlighted the tumoral region.

**Table 4.** Comparison of nnUNet, MTnnUNet, MCnnUNet, AannUNet and Ensemble on rectal tumor segmentation in the external test, fully supervised setting (Num = 666, 9 centers)

| Network | nnUNet | MTnnUNet | MCnnUNet | AA-nnUNet | Ensemble |
| --- | --- | --- | --- | --- | --- |
| aDSC | 62.8 | 65.4 | 60.5 | 65.6 | **69.1** |
| (%) | (60.7, 64.8) | (63.7, 67.2) | (58.6, 62.4) | (63.9, 67.3) | **(67.6, 70.6)** |
| mDSC | 73.2 | 74.4 | 70.2 | 72.8 | **75.5** |
| (%) | (71.8, 74.7) | (73.4, 75.5) | (69.0, 71.5) | (72.0, 73.7) | **(74.5, 76.5)** |
| aHD | 17.28 | **13.63** | 21.06 | 16.69 | 14.74 |
| (mm) | (14.63, 19.94) | **(11.87, 15.38)** | (19.51, 22.62) | (14.57, 18.82) | (12.72, 16.75) |
| mHD | 6.36 | 5.61 | 7.99 | 5.61 | **5.09** |
| (mm) | (5.45, 7.22) | (5.04, 6.18) | (7.00, 9.02) | (4.96, 6.25) | **(4.64, 5.55)** |

aDSC: average Dice Coefficient Similarity. mDSC: median Dice Coefficient Similarity. aHD: average of 95% Hausdorff Distance. mHD: median of 95% Hausdorff Distance. The 95% confidential intervals are presented.

### 4.7 Semi-supervised MTnnUNet, MCnnUNet and AAnnUNet

In the semi-supervised setting, AI-generated pseudo rectum and mesorectum segmentation masks were incorporated during training using training cohort 2, where 141 cases from one center were included for MTnnUNet, MCnnUNet, and AAnnUNet. In the internal validation set, we observed that MTnnUNet showed overall the best results. However, AAnnUNet demonstrated the best results in the external test. In contrast to the purely supervised experiments, MCnnUNet showed better aDSC and aHD in the external set. The reason could be that MCnnUNet was trained and inferred using AI-generated pseudo rectum and mesorectum priors, showing more consistency in performance between internal validation and external test, as shown in Table 5, Table S9, Figure 7b.

**Table 5.** Comparison of nnUNet, MTnnUNet, MCnnUNet, AAnnUNet and Ensemble on rectal tumor segmentation in the external test, semi-supervised setting (Num = 564, 8 centers)

| Network | nnUNet | MTnnUNet | MCnnUNet | AAnnUNet | Ensemble |
| --- | --- | --- | --- | --- | --- |
| aDSC | 66.5 | 69.1 | 70.1 | 71.0 | **71.4** |
| (%) | (64.7, 68.3) | (67.5, 70.6) | (68.7, 71.6) | (69.6, 72.3) | **(70.0, 72.7)** |
| mDSC | 74.4 | 74.9 | 75.7 | 75.8 | **76.3** |
| (%) | (73.1, 75.7) | (73.9, 75.9) | (74.8, 76.6) | (75.0, 76.6) | **(75.4, 77.2)** |
| aHD | 33.84 | 18.55 | 13.26 | **13.14** | 17.66 |
| (mm) | (26.76, 40.93) | (14.52, 22.59) | (11.24, 15.29) | **(10.75, 15.53)** | (13.73, 21.59) |
| mHD | 5.68 | 4.89 | 5.00 | 4.97 | **4.57** |
| (mm) | (5.00, 6.35) | (4.16, 5.62) | (4.30, 5.70) | (4.24, 5.70) | **(4.04, 5.10)** |

aDSC: average Dice Coefficient Similarity. mDSC: median Dice Coefficient Similarity. aHD: average of 95% Hausdorff Distance. mHD: median of 95% Hausdorff Distance. The 95% confidential intervals are presented.

In both supervised and semi-supervised settings, we observed that MTnnUNet exhibited better tumor segmentation results than the baseline model. MCnnUNet demonstrated improvement in the semi-supervised setting, a result that was not observed in the supervised setting. More importantly, the fusion of anomaly maps in the segmentation model ensures more precise tumor localization in both settings. In the first row of Figure S2, nnUNet mistook part of the bladder as the tumor region due to the similar intensity values, and with information fusion from either anatomical masks or anomaly maps, MTnnUNet, MCnnUNet, and AAnnUNet correctly classified the tumor regions.

## 5. Discussion

In this study, we successfully developed a rectal anomaly detection model by training the anatomical inpainting model using prostate T2WI. The model was then applied to the rectal T2WI to generate pseudo-healthy rectal strucures and the anomaly map was the differences between the pseudo-healthy and origial rectal structures. The derived anomaly maps were then used in the downstream tumor segmentation tasks and outperformed the baseline nnUNet, MTnnUNet, which augmented training tasks with the inclusion of rectum and mesorectum segmentation masks, and MCnnunet including rectum and mesorectum as additional input channels, in both the fully-supervised and the semi-supervised manners. In order to automatically generate the rectum and mesorectum mask, we trained an nnUNet to effectively delineate anatomical structures including the normal rectum (without tumoral involvement) and mesorectum.

Research in medical image analysis using AI bears many promises to improve patients’ health. However, Varoquaux et al. (Varoquaux and Cheplygina, 2022) have pointed out that in academia, even though the goal is to solve scientific problems, the emphasis on publication quantity is influenced by Goodhart’s law (Teney et al., 2020), which can compromise scientific quality.

Researchers, in pursuit of novelty, may introduce unnecessary complexity in methods, contributing to technical debt without substantial improvement in predictions. (Isensee et al., 2024) has conducted an extensive benchmarking of current segmentation methods across different dataset and their results revealed a concerning trend that most models introduced in recent years fail to outperform the nnUNet. This is consistent with the findings of this study. Recently published models with higher complexity did not exhibit higher tumor segmentation performance or better generalization ability. Although ViT and SSMs have demonstrated promising results in the natural image classification (Xiao et al., 2023), in rectal tumor contouring using T2WI, transformer-based networks or SSMs-inspired networks do not show better results than CNN-based architectures. The reasons could be, that the transformer relies heavily on large-scale training and remains inferior to CNNs when training data is scarce. Unlike natural images, medical datasets are smaller, typically in the hundreds or thousands (Willemink et al., 2020). Secondly, the quadratic complexity of self-attention poses challenges when dealing with long token sequences (Hatamizadeh et al., 2022a), especially for 3D T2WI.

Beyond architectural optimization, a profound comprehension of medical imaging is integral to advancing tumor segmentation. The rectum and mesorectum provide crucial anatomical context for the localization and delineation of tumors. Several studies (Defeudis et al., 2022; Jayaprakasam et al., 2022) have proved that mesorectal fat and tumor neighboring regions contain important prognostic information in patients with rectal cancer. Therefore, there is a substantial demand for a rectum and mesorectum segmentation model that is accurate and robust. While previous studies (DeSilvio et al., 2023; Hamabe et al., 2022; Kim et al., 2019; Lee et al., 2019) have proposed segmentation networks for these structures, an extensive multi-center external test is highly desirable, and the majority employed 2D models. In this study, we externally tested the anatomical 3D nnUNet and observed successful rectal structure contouring.

Currently, most rectal tumor segmentation studies rely on retrospective datasets, which do not provide a healthy representation of the rectum. Abdominal imaging, particularly prostate MRI, shares overlapping anatomical structures with rectal MRI, and most prostate MRIs display healthy rectal structures. As a cross-domain application, we utilized a public prostate dataset to train the inpainting model, allowing it to learn the distribution of a healthy rectum and mesorectum and generate rectal anomaly maps. Compared to simpler anatomical fusion approaches like MCnnUNet and MTnnUNet, AAnnUNet demonstrated a deeper understanding of rectal and mesorectal anatomy, resulting in improved tumor localization. The anomaly maps also have the potential for other clinical downstream tasks, like lymph node diagnosis, treatment response prediction, and staging stratification.

There are some limitations of this study. First, all the T2WI was annotated by one radiologist. Multiple readers would add extra value to our research. Second, this study exclusively included T2W-MRI, with scans from 2011 to 2018. Some of them exhibited relatively low resolution and larger slice thicknesses, which would not meet the high-resolution criteria established by current protocol recommendations, see Table S1. Third, other MRI sequences like DWI and ADC can potentially improve the segmentation performance. Dou et al. (Dou et al., 2023) have demonstrated by combining T2WI, ADC, and DWI, their model showed the best tumor segmentation performance. Fourth, although we have a relatively large and heterogeneous cohort, the data was solely from the Netherlands. Fifth, concerning rectal anatomical structures, the segmentation process focused solely on the rectum and mesorectum. In future studies, other anatomical structures such as the lumen could contribute to the performance. Lastly, the training and testing were performed exclusively on pre-treatment T2WI. The rectal environment in pre-treatment MRI is visually and pathologically distinct from post-treatment MRI. Incorporating post-treatment T2WI can enhance tumor characterization and improve downstream analytical workflow.

## 6. Conclusion

In this study, we proposed an anatomical inpainting model using prostate MRI and generated rectal anomaly maps from the difference between the original rectal images and reconstructed pseudo healthy slices. We observed that rectal anomaly maps were highly associated with the tumoral areas. To explore the effect of these anomaly maps in the downstream tasks, they were fused as additional input in the rectal tumor segmentation task (AAnnUNet). Other manners of anatomical knowledge fusion including MTnnUNet and MCnnUNet in tumor segmentation were compared. As part of this research, we benchmarked nine DL models as baselines, including CNN-based, transformer-based, and Mamba-based architectures, for rectal tumor segmentation on a large multicenter dataset. nnUNet achieved the best results in the external test set, despite having a relatively lower model complexity compared to the other models, indicating that increased model complexity does not guarantee improved performance. By fusing anatomical knowledge into the tumor segmentation model, MTnnUNet, MCnnUNet, and AAnnUNet demonstrated improved performance compared to nnUNet. Among these, AAnnUNet achieved the highest accuracy in rectal tumor segmentation. Anomaly map generation requires segmentation masks of the rectum and mesorectum. We trained an nnUNet for this, achieving high accuracy with an aDSC over 0.80 on the external dataset. MTnnUNet, MCnnUNet, and AAnnUNet, when trained with AI-generated pseudo rectum and mesorectum masks, significantly improved rectal tumor segmentation performance compared to the baseline model, with AAnnUNet achieving the best results in tumor segmentation. Our analysis indicates that integrating anomaly maps derived from inpainting can effectively enhance tumor segmentation performance. Additionally, the anomaly maps, which are strongly associated with rectal tumors, can be utilized in other downstream tasks to actively monitor rectal cancer. The code, model weights and the manually annotated rectum masks for prostate T2WI and can be accessed here: https://github.com/Liiiii2101/Anatomy-aware-nnUNet-for-Rectal-Tumor-Segmentation.

## Data Availability

All code, trained weights and annotated prostate rectum and mesorectum masks will be made publicly available upon publication upon https://github.com/Liiiii2101/Anatomy-aware-nnUNet-for-Rectal-Tumor-Segmentation. The original data of rectal cancer patient is private and is not publicly available to guarantee protection of patient privacy.

## Acknowledgment

The study was supported by the Research High Performance Computing (RHPC) facility of the Netherlands Cancer Institute.

## Funding sources

This study has received funding from the European Union’s Horizon 2020 Research and Innovation Programme under the Marie Skłodowska-Curie grant agreement No 857894.

## Notes

### Competing Interest Statement

The authors have declared no competing interest.

### Funding Statement

This project has received funding from the European Union Horizon 2020 Research and Innovation Programme under the Marie Sklodowska-Curie grant agreement No 857894.

### Author Declarations

The study was conducted in accordance with the Declaration of Helsinki and has been approved by the Institutional Review Board (IRB) of the Netherlands Cancer Institute. Each participating centre reviewed the study protocol and provided approval. Informed consent was waived by the IRB and by each participating centre during local ethical review and approval due to the retrospective nature of the study.

