## Supplementary for "Improving Rectal Tumor Segmentation with Anomaly Fusion Derived from Anatomical Inpainting: A Multicenter Study"

### Supplemental Materials

#### Table S1

#### Acqusition Parameters

| | Vendor | Model | Field Strength (T) | TR (ms) | TE (ms) | Flip Angle (degrees) | NSA | Slice Thickness (mm) | Pixel Spacing (mm) | FOV (mm) |
|---|---|---|---|---|---|---|---|---|---|---|
| **Center 1** | Siemens | Aera | 1.5 | 3620-4830 | 78-84 | 145-173 | 3 | 3 | 0.35x0.35 | 220x220-230x230 |
| | | Avanto | 1.5 | 3340-5520 | 96-100 | 148-150 | 2-3 | 3.5 | 0.63x0.63 | 200x200 |
| | | Espree | 1.5 | 3000-5860 | 98-102 | 150 | 2-3 | | 0.63x0.63-0.69x0.69 | 200x200-220x220 |
| | | Symphony | 1.5 | 3000 | 68 | 160 | 1 | 4 | 0.63x0.63 | 200x200 |
| **Center 2** | Philips | Ingenia | 1.5 | 2199-4563 | 60 | 90 | 2 | 3.5 | 0.49x0.49 | 250x250 |
| | | Achieva | 1.5 | 3525-3533 | 60 | 90 | 3 | 3.5 | 0.49x0.49 | 250x250 |
| **Center 3** | GE | Signa HDxt | 1.5 | 3880-8528 | 101-109 | 90 | 1.5-2 | 4-4.4 | 0.47x0.47-0.59x0.59 | 240x240-300x300 |
| | | Signa HDxt | 3.0 | 2302-7580 | 82-132 | 90-130 | 1-5 | 3-4.4 | 0.31x0.31-0.47x0.47 | 160x160-240x240 |
| | Philips | Ingenia | 3.0 | 3655-5605 | 100 | 90 | 1 | 3 | 0.47x0.47 | 180x180 |
| | Siemens | Aera | 1.5 | 3680-9030 | 72-101 | 130-160 | 1-3 | 3.5 | 0.63x0.63 | 200x200 |
| | | Avanto | 1.5 | 3390-6370 | 70-84 | 150 | 3-4 | 3-3.5 | 0.78x0.78-0.99x0.99 | 200x200-362x379 |
| **Center 4** | GE | Optima MR450w | 1.5 | 9333-9381 | 115-130 | 160 | 1.5-2 | 4 | 0.47x0.47 | 240x240 |
| | Philips | Ingenia | 1.5 | 2200-3950 | 70 | 90 | 1-2 | 3-4 | 0.29x0.29-0.35x0.35 | 180x180-220x220 |
| | | Intera | 1.5 | 2200-2501 | 70 | 90 | 4 | 3-4 | 0.35x0.35 | 180x180 |
| **Center 5** | Siemens | Avanto | 1.5 | 2550-9220 | 102-122 | 120-150 | 1-2 | 3-3.5 | 0.47x0.47-0.94x0.94 | 240x240-310x310 |
| | | Avanto fit | 1.5 | 3400-6120 | 122 | 133-150 | 2 | 3 | 0.73x0.73-0.78x0.78 | 280x280-300x300 |
| **Center 6** | Philips | Ingenia | 1.5 | 1593-16738 | 90-130 | 90 | 2 | 3 | 0.56x0.56-0.78x0.78 | 180x180-200x200 |
| | | Intera | 1.5 | 2255-11275 | 130 | 90 | 4-6 | 3 | 0.78x0.78 | 200x200 |
| **Center 7** | Siemens | Espree | 1.5 | 3300-8680 | 101-114 | 145-150 | 2-3 | 3-5 | 0.47x0.47-0.88x0.88 | 150x150-280x280 |
| | | SymphonyTim | 1.5 | 3000-6430 | 90-108 | 140-150 | 1-2 | 3-5 | 0.63x0.63-1.48x1.48 | 200x200-380x380 |
| | | Verio | 3.0 | 3540-8590 | 95-101 | 143-150 | 1-2 | 3-5 | 0.63x0.63-0.88x0.88 | 200x200-280x278 |
| **Center 8** | Philips | Achieva | 3.0 | 1573-4250 | 120-150 | 90 | 2-3 | 3 | 0.48x0.48-0.49-0.49 | 230x230-250x250 |
| | | Achieva dStream | 3.0 | 3686-6395 | 120 | 90 | 1 | 3 | 0.39x0.39-0.40x0.40 | 277x277-282x282 |
| | | Ingenia | 3.0 | 7088 | 120 | 90 | 1 | 4 | 0.39x0.39 | 281x281 |
| **Center 9** | Philips | Achieva dStream | 1.5 | 866-4703 | 120-250 | 90 | 1 | 3-5 | 0.63x0.63-1.19x1.19 | 240x240-400x400 |
| | Siemens | Verio | 3.0 | 2520-9390 | 101-163 | 112-150 | 1-2 | 3-4 | 0.54x0.54-0.94x0.94 | 240x240 |

**Table S2**

**Rectum and Mesorectum Annotation Status**

| Center | C1 | C2 | C3 | C4 | C5 | C6 | C7 | C8 | C9 | Total |
|---|---|---|---|---|---|---|---|---|---|---|
| Annotated | 23 | 26 | 20 | 25 | 11 | 11 | 25 | 0 | 39 | 180 |
| Total | 26 | 88 | 20 | 135 | 11 | 110 | 141 | 42 | 132 | 705 |

Note: C, center. Annotated, number of samples with manually annotated rectum and mesorectum. Total, the total number of samples in each center. All samples have manually annotated tumor masks.

**Table S3**

**Comparison of various models on rectal tumor in the 5-fold cross-validation (Num = 39)**

| Network | UNet | ResUNet | UNetR | SwinUNetR | Atten-UNet | MedFormer | nnFormer | U-Mamba | nnUNet |
|---|---|---|---|---|---|---|---|---|---|
| aDSC (%) | 59.0 (52.0, 65.9) | 58.7 (52.1, 65.2) | 48.9 (41.9, 55.9) | 54.6 (47.9, 61.3) | 59.4 (53.1, 65.6) | **66.3** (60.1, 72.5) | 48.2 (38.3, 58.2) | 57.3 (47.9, 66.8) | 63.2 (55.7, 70.7) |
| mDSC (%) | 65.7 (60.1, 71.2) | 63.3 (55.2, 71.4) | 55.3 (44.1, 66.5) | 56.0 (44.9, 67.1) | 65.4 (59.6, 71.2) | 69.5 (61.9, 77.1) | 56.9 (37.8, 76.0) | **72.0** **(61.3, 82.6)** | 70.6 (63.3, 78.0) |
| aHD (mm) | 15.03 (9.74, 20.31) | **14.52** **(10.86, 18.17)** | 47.89 (29.57, 66.21) | 21.00 (15.48, 26.52) | 17.08 (10.97, 23.19) | 15.03 (8.24, 21.82) | 14.84 (8.23, 21.45) | 23.90 (5.70, 42.10) | 23.27 (4.92, 41.63) |
| mHD (mm) | 9.82 (5.30, 14.33) | 12.35 (7.10, 17.60) | 30.70 (17.20, 44.21) | 25.21 (23.05, 27.34) | 8.56 (4.80, 12.32) | **6.39** (2.39, 10.38) | 10.00 (3.14, 16.87) | 7.85 (4.80, 12.32) | 7.19 (3.47,10.90) |

aDSC: averaged Dice Coefficient Similarity. aHD: averaged of 95% Hausdorff Distance. mDSC: median Dice Coefficient Similarity. mHD: median of 95% Hausdorff Distance. The 95% confidential intervals are presented.

**Table S4**

| | UNet | ResUNet | UNetR | SwinUNerR | Atten-UNet | nnFormer | MedFormer | U-Mamba | nnUNet |
|---|---|---|---|---|---|---|---|---|---|
| UNet | - | ns | **** | **** | ns | **** | *** | * | **** |
| ResUNet | | - | **** | **** | ns | **** | *** | * | **** |
| UNetR | | | - | ns | **** | * | **** | **** | **** |
| SwinUNetR | | | | - | **** | ** | **** | **** | **** |
| Atten-UNet | | | | | - | **** | **** | *** | **** |
| nnFormer | | | | | | - | **** | **** | **** |
| MedFormer | | | | | | | - | ns | **** |
| U-Mamba | | | | | | | | - | **** |
| nnUNet | | | | | | | | | - |

DSC, external test set. ns: p > 0.05, *: p <= 0.05, **: p <= 0.01, ***: p <= 0.001, ****: p <= 0.0001

**Table S5**

| | nnUNet | MTnnUNet | MCnnUNet | AAnnUNet | Ensemble |
|---|---|---|---|---|---|
| nnUNet | - | **** | ** | *** | **** |
| MTnnUNet | | - | **** | ns | **** |
| MCnnUNet | | | - | **** | **** |
| AAnnUNet | | | | - | **** |
| Ensemble | | | | | - |

DSC,  trained on training cohort 1 (num=39) external test set (num=666). ns: p > 0.05, *: p <= 0.05, **: p <= 0.01, ***: p <= 0.001, ****: p <= 0.0001

**Table S6**

| | nnUNet | MTnnUNet | MCnnUNet | AAnnUNet | Ensemble |
|---|---|---|---|---|---|
| nnUNet | - | **** | **** | **** | **** |
| MTnnUNet | | - | * | **** | **** |
| MCnnUNet | | | - | * | ** |
| AAnnUNet | | | | - | ns |
| Ensemble | | | | | - |

DSC,  trained on training cohort 1 (num=141) external test set (num=564).. ns: p > 0.05, *: p <= 0.05, **: p <= 0.01, ***: p <= 0.001, ****: p <= 0.0001

**Table S7**

**The performance of rectum and mesorectum segmentation using nnUNet in the external cohort (Num = 141, 7 centers)**

| Structures | Rectum | Mesorectum |
|---|---|---|
| aDSC (%) | 87.3 (85.6, 88.9) | 81.4 (79.6, 83.2) |
| mDSC (%) | 90.0 (89.3, 90.7) | 84.2 (83.2, 85.2) |
| aHD(mm) | 10.15 (7.33, 12.97) | 10.57 (3.70, 17.44) |
| mHD (mm) | 3.19 (1.70, 4.68) | 4.39 (3.64, 5.15) |

aDSC: averaged Dice Coefficient Similarity. aHD: averaged of 95% Hausdorff Distance. mDSC: median Dice Coefficient Similarity. mHD: median of 95% Hausdorff Distance. The 95% confidential intervals are presented.

**Table S8**

**nnUNet vs. MTnnUNet vs. MCnnUNet vs. AAnnUNet on rectal tumor segmentation in the internal validation (Num = 39, 1 center, 5-fold cross-validation)**

| Network | nnUNet | MTnnUNet | MCnnUNet | AAnnUNet |
|---|---|---|---|---|
| aDSC (%) | 63.2 (55.7, 70.7) | 64.8 (57.6, 72.0) | **73.2 (66.1, 80.3)** | 68.7 (62.5, 74.8) |
| mDSC (%) | 70.6 (63.3, 78.0) | 70.2 (67.2, 73.2) | **80.8 (78.3, 83.3)** | 74.0 (68.6, 79.3) |
| aHD (mm) | 23.27 (4.92, 41.63) | 10.98 (7.36, 14.60) | **5.86 (3.44, 8.28)** | 8.85 (5.60, 12.09) |
| mHD (mm) | 7.19 (3.45, 10.90) | 6.23 (3.87, 8.59) | **3.45 (2.65, 4.35)** | 4.38 (2.53, 6.22) |

aDSC: averaged Dice Coefficient Similarity. aHD: averaged of 95% Hausdorff Distance. mDSC: median Dice Coefficient Similarity. mHD: median of 95% Hausdorff Distance. The 95% confidential intervals are presented.

**Table S9**

**nnUNet vs. MTnnUNet vs. MCnnUNet vs. AAnnUNet on rectal tumor segmentation in the internal validation (Num = 141, 1 center, 5-fold cross-validation)**

| Network | nnUNet | MTnnUNet | MCnnUNet | AAnnUNet |
|---|---|---|---|---|
| aDSC (%) | 72.3 (69.1, 75.4) | **72.7 (69.9, 75.6)** | 71.9 (68.9, 74.9) | 71.8 (68.8, 74.8) |
| mDSC (%) | **78.7 (77.1, 80.3)** | 78.6 (75.8, 81.4) | 78.1 (76.4, 79.8) | 77.0 (75.4, 78.5) |
| aHD (mm) | 10.55 (7.35, 13.74) | **9.01 (6.78, 11.38)** | 11.40 (7.82, 14.99) | 10.93 (7.52, 14.35) |
| mHD (mm) | **3.91 (3.04, 4.77)** | **3.91 (3.00, 4.81)** | **3.91 (2.84, 4.96)** | **3.91 (3.00, 4.81)** |

aDSC: averaged Dice Coefficient Similarity. aHD: averaged of 95% Hausdorff Distance. mDSC: median Dice Coefficient Similarity. mHD: median of 95% Hausdorff Distance. The 95% confidential intervals are presented.

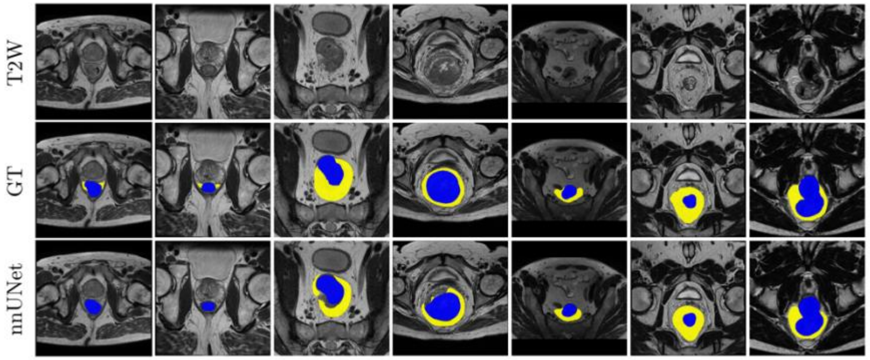


**Figure S1** The visualization of the segmentation performance of nnUNet for rectum (blue) and mesorectum (yellow). Each column is a different sample from external set. The rows from top to bottom are: original T2WI, ground truth, and predictions from nnUNet.

**Table S10**

**Comparison of various models on rectal tumor segmentation in the 5-fold cross-validation (Num = 132, center 9 only , 5-fold cross-validation)**

| Network | UNet | ResUNet | UNetR | SwinUNetR | Atten-UNet | MedFormer | nnFormer | U-Mamba | nnUNet |
|---|---|---|---|---|---|---|---|---|---|
| aDSC (%) | 71.4 (68.9, 74.0) | 72.1 (69.5, 74.8) | 55.8 (52.3, 59.3) | 59.7 (56.7, 62.8) | 71.3 (68.3, 74.3) | 70.2 (67.1, 73.2) | 69.4 (65.6, 73.1) | **73.5 (70.2, 76.7)** | **73.5 (70.1, 77.0)** |
| mDSC (%) | 75.5 (73.7, 77.3) | 77.7 (75.8, 79.7) | 60.9 (57.3, 64.5) | 64.6 (61.7, 67.4) | 75.8 (73.9, 77.7) | 75.4 (73.9, 76.9) | 76.4 (73.8, 78.9) | 78.4 (75.9, 80.9) | **79.6 (77.6, 81.6)** |
| aHD (mm) | 11.60 (8.28, 14.90) | **9.14 (7.29, 10.99)** | 34.68 (27.12, 42.24) | 21.13 (17.4, 24.9) | 11.69 (8.61, 14.76) | 14.03 (10.26, 17.80) | 12.72 (9.08, 16.36) | 9.90 (4.27,15.54) | 12.20 (5.46, 18.93) |
| mHD (mm) | 4.43 (3.68, 5.18) | 4.67 (3.43, 5.90) | 14.56 (8.24, 20.87) | 13.92 (9.73, 18.11) | 4.61 (3.53, 5.69) | 5.10 (3.49, 6.72) | 5.21 (3.65, 6.76) | **3.34 (2.88, 3.79)** | 3.64 (3.13, 4.17) |

aDSC: averaged Dice Coefficient Similarity. aHD: averaged of 95% Hausdorff Distance. mDSC: median Dice Coefficient Similarity. mHD: median of 95% Hausdorff Distance. The 95% confidential intervals are presented.

**Table S11**

**Comparison of various models on rectal tumor segmentation in the external test (Num = 573, 8 centers)**

| Network | UNet | ResUNet | UNetR | SwinUNetR | Atten-UNet | MedFormer | nnFormer | U-Mamba | nnUNet |
|---|---|---|---|---|---|---|---|---|---|
| aDSC (%) | 68.9 (67.2, 70.5) | 69.5 (67.9, 71.2) | 49.8 (47.7, 51.8) | 52.0 (49.9, 54.0) | 70.2 (68.6, 71.7) | 69.8 (68.0, 71.4) | 63.4 (61.2, 65.6) | 70.6 (68.7, 72.4) | **72.4 (70.6, 74.2)** |
| mDSC (%) | 75.8 (74.8, 76.9) | 75.9 (74.6, 77.1) | 55.5 (53.2, 57.8) | 59.3 (56.8, 61.8) | 76.1 (75.1, 77.0) | 76.3 (75.3, 77.3) | 74.4 (73.0, 75.8) | 78.7 (77.4, 80.0) | **79.7 (78.7, 80.7)** |
| aHD (mm) | 18.32 (15.21,21.42) | 15.65 (13.19, 18.12) | 37.30 (32.21, 42.39) | 28.57 (25.52, 31.61) | 15.46 (12.90,18.01) | 15.53 (13.15, 17.90) | 21.33 (17.63, 25.03) | 12.84 (10.20, 15.49) | **11.59 (9.31, 13.86)** |
| mHD (mm) | 5.00 (4.52, 5.48) | 4.65 (4.20, 5.10) | 14.04 (12.24, 15.85) | 13.94 (12.57,15.31) | 4.76 (4.23, 5.28) | 4.75 (4.32, 5.18) | 6.12 (5.40, 6.84) | 3.91 (3.38, 4.43) | **3.64 (3.26, 4.03)** |

aDSC: averaged Dice Coefficient Similarity. aHD: averaged of 95% Hausdorff Distance. mDSC: median Dice Coefficient Similarity. mHD: median of 95% Hausdorff Distance. The 95% confidential intervals are presented.

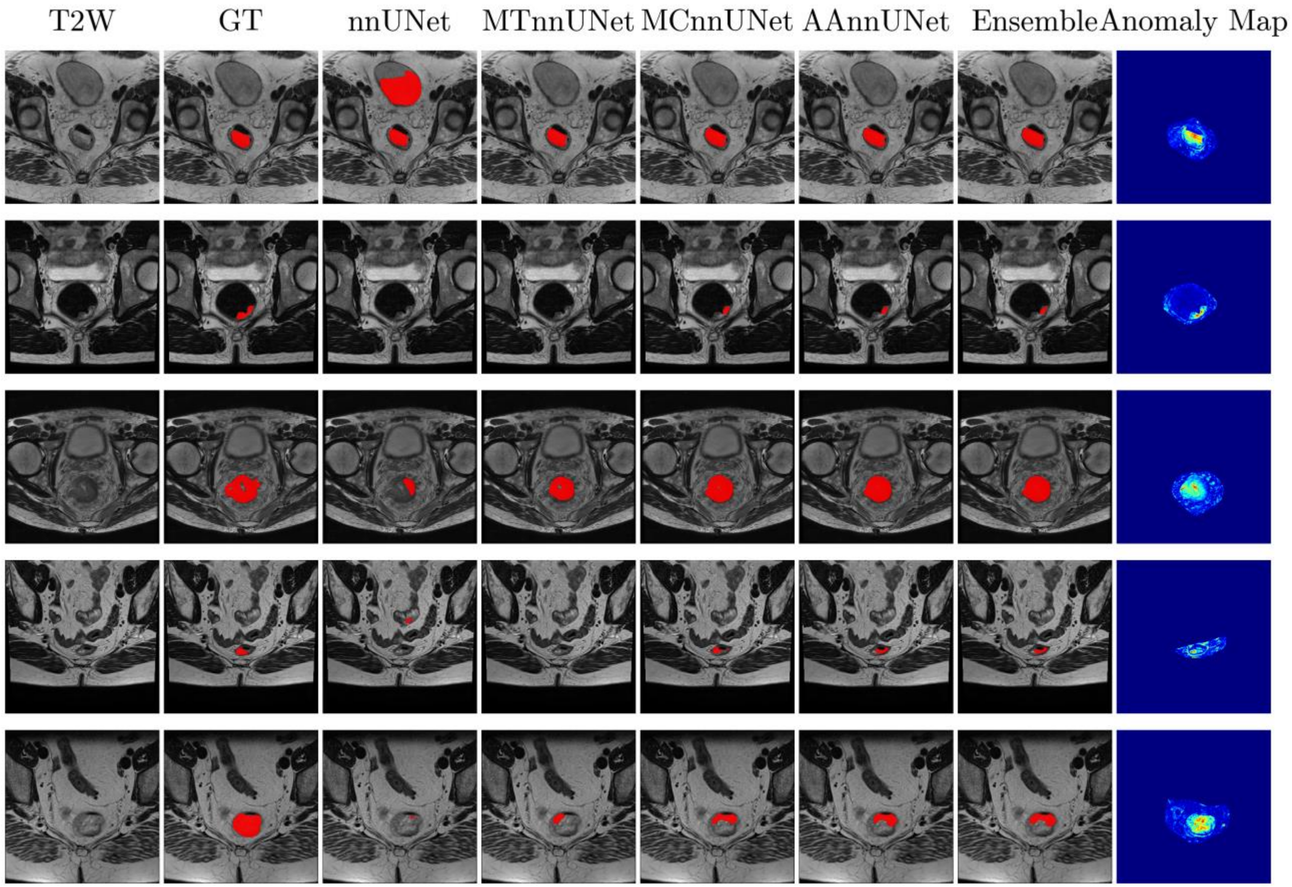


**Figure S2** The visualization of the segmentation performance of nnUNet, MTnnUNe, MCnnUNet, AAnnUNet and ensemble using T2WI from different centers, semi-supervised setting. Each row is a different sample. The columns from left to right are original T2WI, ground truth, tumor prediction mask from nnUNet, MTnnUNet, MCnnUNet, AAnnUNet, and Ensemble.

**Metrics:**

Dice Similarity Coefficient (DSC)

$$DSC = \frac{2P \cap G}{|P| + |G|}$$

Hausdorff Distance:

$$d(P,G)_H = max\{sup_{p \in P}\, inf_{g \in G}\, d(p,g), sup_{p \in P}\, inf_{g \in G}\, d(g,p)\}$$

Where *P* represents the predicted segmentation mask, and *G* is the ground truth. If DSC equals 1, it indicates a perfect predicted segmentation. *d*(·) denotes the Euclidean distance. sup and *inf* are supremum and infimum. In our study, to levitate the impact of the outliers, 95% HD was utilized.

Structural Similarity Index Measurement (SSIM),

$$SSIM = \frac{\left(2\mu_{y(x)}\mu_{G(x)} + c_1\right)\left(2\sigma_{y(x)G(x)} + c_2\right)}{\left(\mu_{y(x)}^2 + \mu_{G(x)}^2 + c_1\right)\left(\sigma_{y(x)}^2 + \sigma_{G(x)}^2 + c_2\right)}$$

Peak Signal-to-Noise Ratio (PSNR)

$$PSNR = 10 \log_{10} \frac{\overset{2}{\max}\left(y(x), G(x)\right)}{\frac{1}{N}|y(x) - G(x)|_2^2}$$

where $G(x)$ is a generated image, $y(x)$ is a ground-truth image, $\mu_{y(x)}$ and $\mu_{G(x)}$ denotes the mean of $y(x)$ and $G(x)$, respectively, $\sigma_{y(x)}$ and $\sigma_{G(x)}$ represent the variance of $y(x)$ and $G(x)$, respectively, $\sigma_{y(x)G(x)}$ is the covariance of $y(x)$ and $G(x)$, and $c_1$ and $c_2$ represent positive constants used to avoid null denominators.

Preprocessing Details:

**Train with training cohort 1,**

Target sampling spacing: [2.9999978065490724, 0.7291666865348816, 0.7291666865348816]

Patch size: [32, 256, 224]

**Train with training cohort 2,**

Target sampling spacing: [4.999992847442627, 0.78125, 0.78125]

Patch size: [28, 256, 256]

For more models' summary and code, see: https://github.com/Liiiii2101/Anatomy-aware-nnUNet-for-Rectal-Tumor-Segmentation
